# Detecting somatic variants in purified brain DNA obtained from surgically implanted depth electrodes in epilepsy

**DOI:** 10.1101/2024.07.08.24310005

**Authors:** Rumika Mascarenhas, Daria Merrikh, Maryam Khanbabaei, Navprabhjot Kaur, Navid Ghaderi, Tatiana Maroilley, Yiping Liu, Tyler Soule, Juan Pablo Appendino, Julia Jacobs, Samuel Wiebe, The Calgary Comprehensive Epilepsy Program Collaborators, Walter Hader, Gerald Pfeffer, Maja Tarailo-Graovac, Karl Martin Klein

**Affiliations:** Department of Clinical Neurosciences, University of Calgary, Calgary, AB, Canada; Alberta Children’s Hospital Research Institute, University of Calgary, Calgary, AB, Canada; Hotchkiss Brain Institute, University of Calgary, Calgary, AB, Canada; Department of Medical Genetics, Cumming School of Medicine, University of Calgary, Calgary, AB, Canada; Department of Biochemistry and Molecular Biology, Cumming School of Medicine, University of Calgary, Calgary, AB, Canada; Flow Cytometry Core Facility, University of Calgary, Calgary, AB, Canada; Department of Pediatrics, University of Calgary, Alberta Children’s Hospital, Calgary, AB, Canada; Department of Community Health Sciences, University of Calgary, Calgary, AB, Canada; O’Brien Institute for Public Health, University of Calgary, Calgary, AB, Canada; Clinical Research Unit, Cumming School of Medicine, University of Calgary, Calgary, AB, Canada

## Abstract

Somatic variants causing epilepsy are challenging to detect as they are only present in a subset of brain cells (e.g., mosaic) resulting in low variant allele frequencies. Traditional methods that rely on surgically resected brain tissue are limited to patients undergoing brain surgery. We developed an improved protocol to detect somatic variants using DNA from stereo- electroencephalography (SEEG) depth electrodes, enabling access to a larger patient cohort and diverse brain regions. This protocol mitigates issues of cell contamination and low yields by purifying neuronal nuclei using fluorescence-activated nuclei sorting. Furthermore, we employed advanced amplification techniques, stringent quality control and an optimized bioinformatic workflow to decrease false positives. Using digital droplet polymerase chain reaction, we confirmed all four selected candidate somatic variants. Our approach enhances the reliability and applicability of SEEG-derived DNA for epilepsy, offering insights into its molecular basis, facilitating identification of the epileptogenic zone and other advancements in precision medicine.

## Introduction

Brain somatic variants are increasingly recognized as a cause of epilepsy, particularly of focal epilepsies caused by brain malformations^1–11^. Detecting these variants is crucial to understanding the processes underlying the generation of epileptic seizures and, consequently, improving treatment options. However, detection of somatic variants in the human brain is challenging as they are only present in a subset of cells in the affected portion of the brain.

Traditionally, brain tissue for detection of somatic variants is obtained from patients undergoing resective epilepsy surgery. While this method successfully identifies somatic variants in several types of lesional epilepsies^1–11^, it bears limitations. One such limitation is that only a small proportion of patients with epilepsy are eligible for resective surgery that provides tissue for analysis, which means only a subset of patients are investigated. This causes a bias towards the identification of somatic variants in patients with surgically treatable epilepsy, who often have underlying brain lesions such as brain malformations. Additionally, even in patients undergoing resective surgery, access to the unaffected tissue is limited, which restricts the extent of comparative analyses. Furthermore, in patients with non-lesional epilepsies, it is often unclear which part of the tissue should be selected to obtain DNA for identification of somatic variants.

Given the above limitations and the importance of exploring somatic brain variants beyond surgical candidates, there is an urgent need for novel methods to examine brain-derived DNA in patients with focal epilepsies. Recently, a novel approach of DNA extraction has been proposed which involves examining trace DNA obtained from depth electrodes used for stereo- electroencephalography (SEEG) to detect somatic brain variants associated with focal epilepsy^12–15^. The SEEG procedure uses depth electrodes surgically implanted into the brain through skull burr holes to record seizure activity and determine the source of the seizures for possible resection.

Harvesting brain somatic DNA from SEEG electrodes allows inclusion of patients who do not eventually undergo epilepsy surgery and access to brain tissue from multiple regions where electrodes are implanted. The spatial distribution of the identified somatic variants can then be correlated with the seizure onset zone allowing prioritization of variants only present in the seizure onset zone.

## Design

The originally proposed SEEG harvesting method has significant shortcomings. Firstly, the pool of cells that is obtained from the depth electrodes can contain contaminants, such as blood, or immune cells, in addition to brain cells. The presence of cell types other than neurons lowers the percentage of brain cells containing the variant, resulting in lower variant allele frequencies (VAFs) as observed in prior studies^14,15^. This makes it more challenging to detect disease-causing somatic variants. Secondly, compared to resected brain tissue, the number of cells obtained through this method is relatively low, necessitating the amplification of DNA to obtain sufficient amounts for sequencing. This process introduces amplification artifacts in the samples, which can lead to false positive findings in downstream variant calling processes.

Here, we describe an improved protocol outlining our optimized SEEG harvesting technique combined with deep exome sequencing and computation analyses which successfully addresses the aforementioned limitations. Using this protocol, we identified a somatic variant in *MTOR* at a VAF of 0.78% in a patient with focal cortical dysplasia (FCD)^16^. Our protocol, described herein 1) introduces purification of neuronal nuclei from SEEG electrodes addressing the issue of cell contamination and uses a novel amplification method^17^ which reduces amplification artifacts; 2) introduces pre- and post- sequencing quality control steps that allow selection of high-quality samples, and 3) outlines a bioinformatic variant filtration workflow for reliable somatic variant detection. An overview of our methodology is shown in Fig.1.

**Fig. 1:**
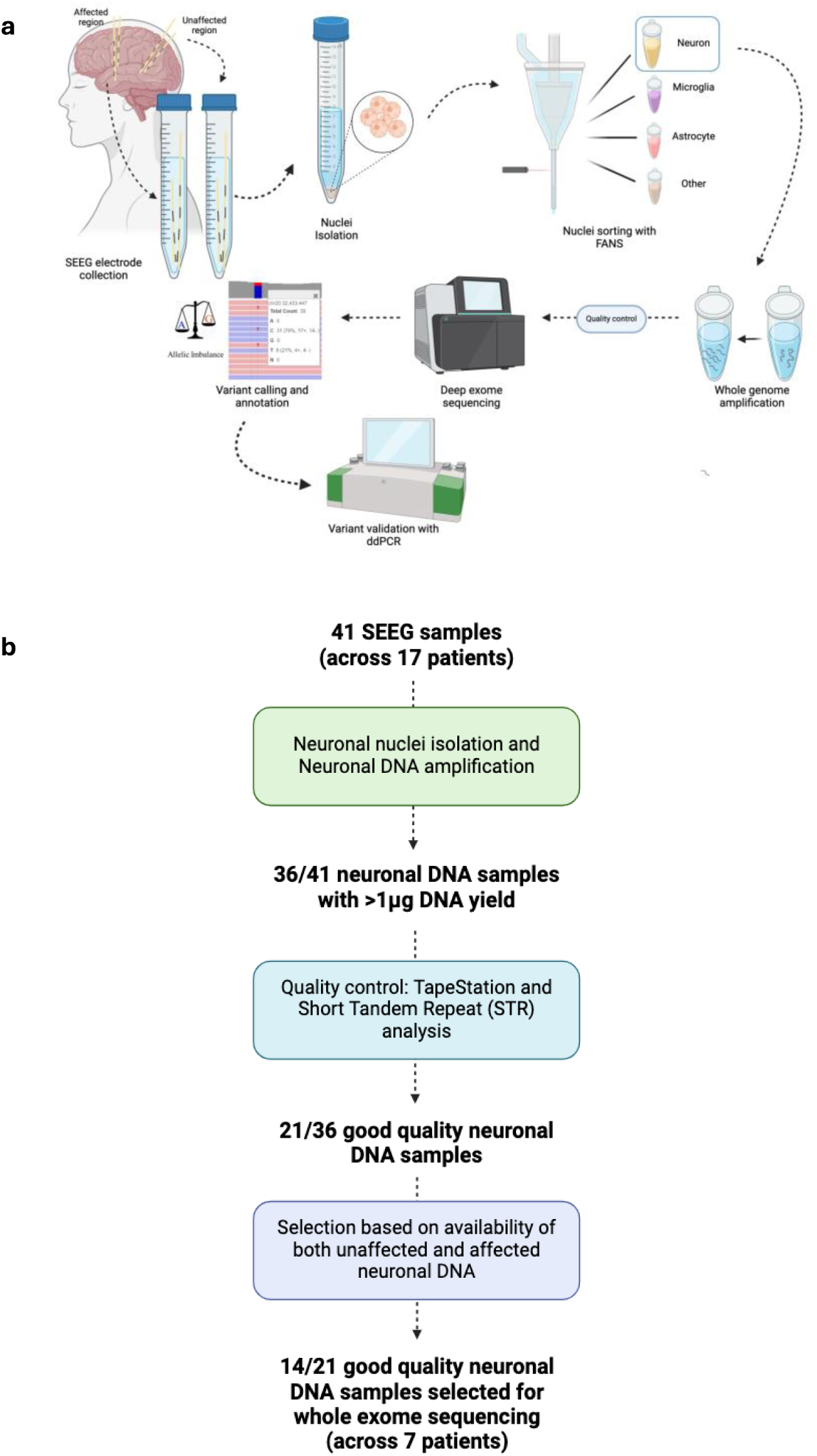
Overview of methodology for SEEG-derived DNA analysis to detect somatic brain variants. a, Workflow summarizing the steps involved in DNA extraction from neuronal nuclei isolated from SEEG depth electrodes to detect somatic brain variants. **b,** Flowchart showing how SEEG samples were selected for whole exome sequencing.

## Results

### Neuronal nuclei isolation using fluorescence-activated nuclei sorting (FANS) and DNA amplification using primary template amplification (PTA)

To avoid inclusion of contaminating cells, our protocol uses FANS to isolate nuclei and purify neuronal nuclei. Our protocol is based on Nott et al.^18^ with modifications to accommodate for differences associated with processing of depth electrodes in comparison to brain tissue and avoid fixation for optimal DNA quality. We use 4′,6-diamidino-2-phenylindole (DAPI) to stain nuclei containing DNA and NeuN to stain neuronal nuclei. To optimize the sorting process, we also stain microglia (SPI1) and astrocyte nuclei (LHX2). We modified the gating strategy used for FANS to increase nuclei yield and quality (see Methods).

We processed depth electrodes from 41 brain regions (17 affected and 24 unaffected) across 17 patients with focal epilepsy. On average, three electrodes were pooled per region (range 1-5). Affected brain regions were selected based on the seizure onset recorded on SEEG and/or presence of a lesion on MRI. Unaffected regions were selected to be most distant from the affected brain MRI lesion and least involved on SEEG. Depth electrodes from affected and unaffected brain regions were processed similarly. Our protocol yielded an average of 713 neuronal nuclei per region (Table 1), representing on average 6% of the unsorted sample, which were purified using FANS. 80% of the samples had ≥112 neuronal nuclei (90% ≥40) corresponding to 80% power to identify a VAF of ≥0.7% (2.0%).

**Table 1:** Number of nuclei per pool for each cell type harvested from SEEG electrodes using our optimized method. . 41 brain regions across 17 patients. Triple negative nuclei were DAPI- positive indicating the presence of DNA but were negative for neuron, astrocyte and microglia markers.

| Nuclei Type | Average | Median | Range |
| --- | --- | --- | --- |
| Neuron | 713 | 225 | 12-6,424 |
| Astrocyte | 1,104 | 70 | 2-12,463 |
| Microglia | 1,776 | 132 | 6-43,644 |
| Triple Negative | 37,943 | 19,732 | 108-208,402 |

DNA from sorted neuronal nuclei was amplified using PTA, a newly published method that guides the polymerase to rebind with the original DNA strand. This ensures more uniform genome amplification and minimizes errors, resulting in enhanced variant detection accuracy and sensitivity^17,19^. Overall, 89% (36/41) of the amplified SEEG-derived neuronal DNA samples had >1 *µ*g DNA yield in total, on which further quality control was performed.

### Short tandem repeat (STR) analysis can be used to exclude low quality samples prior to whole exome sequencing (WES)

Amplified SEEG-derived neuronal DNA samples with sufficient DNA yield (>1 *µ*g) for sequencing (36/41) were subjected to two quality control measures: (1) Fragment size analysis using TapeStation, to determine accurate sizes of DNA fragments and to ensure high quality of the samples and (2) STR analysis to confirm the presence of identical human DNA in the amplified SEEG-derived neuronal DNA.

For fragment size analysis, the distribution of the DNA fragment sizes in the samples were analysed. Peak sizes in the results indicate the presence of the dominant fragment size in the respective sample. The average peak size of the DNA fragments across the SEEG-derived neuronal DNA samples with DNA yield >1 *µ*g (36 samples) was 1338bp and ranged from 1007bp to 1548bp, which is in the expected range for PTA (200 – 4000bp).

For STR analysis, the pattern of alleles in 15 STR and 1 sex markers in the 36 amplified SEEG-derived neuronal DNA samples were compared to the pattern of alleles in their corresponding blood- or saliva-derived DNA samples. The markers are spread across the genome, and absence of these markers suggests uneven amplification across the genome. The number of markers with allelic dropout were used as a measure for STR analysis quality. Allelic dropout was described if a STR or sex marker was missing or if only one allele was present in the neuronal samples while two alleles were present in the germline (blood/saliva) sample. We categorized the samples based on their quality as interpreted from their STR analysis results. 58% (21/36) of the SEEG-derived neuronal DNA samples had ≤4 markers with allelic dropout which corresponded to 9/17 patients (53%).

For our WES run, we chose the best patient samples, as determined by a complete set of good quality affected SEEG-derived neuronal DNA, unaffected SEEG-derived neuronal DNA and germline (blood/saliva-derived) DNA samples. Consequently, 14/21 SEEG-derived neuronal DNA samples (7 affected and 7 unaffected) across 7 patients were selected (Supplementary Table 1). The quality of the 14 SEEG-derived neuronal DNA samples on the STR analysis varied between 0 to 3 markers with allelic dropout (Supplementary Table 2).

### Exome coverage following sequencing

Following sequencing and alignment of sequences to the reference genome, we analyzed the post-alignment quality of the samples using Qualimap^20^. Among the 14 SEEG-derived neuronal DNA samples analyzed, 10 samples exhibited good coverage, with ≥90% of the exome being covered by at least 95 reads (Fig. 2a). In these 10 samples, the STR analysis results demonstrated ≤ 1 STR marker with allelic dropout (Supplementary Table 2). Conversely, the remaining 4 samples displayed lower coverage, with 60-79% of the exome covered at a minimum of 95 reads corresponding to higher allelic dropout on STR analysis.

**Fig. 2:**
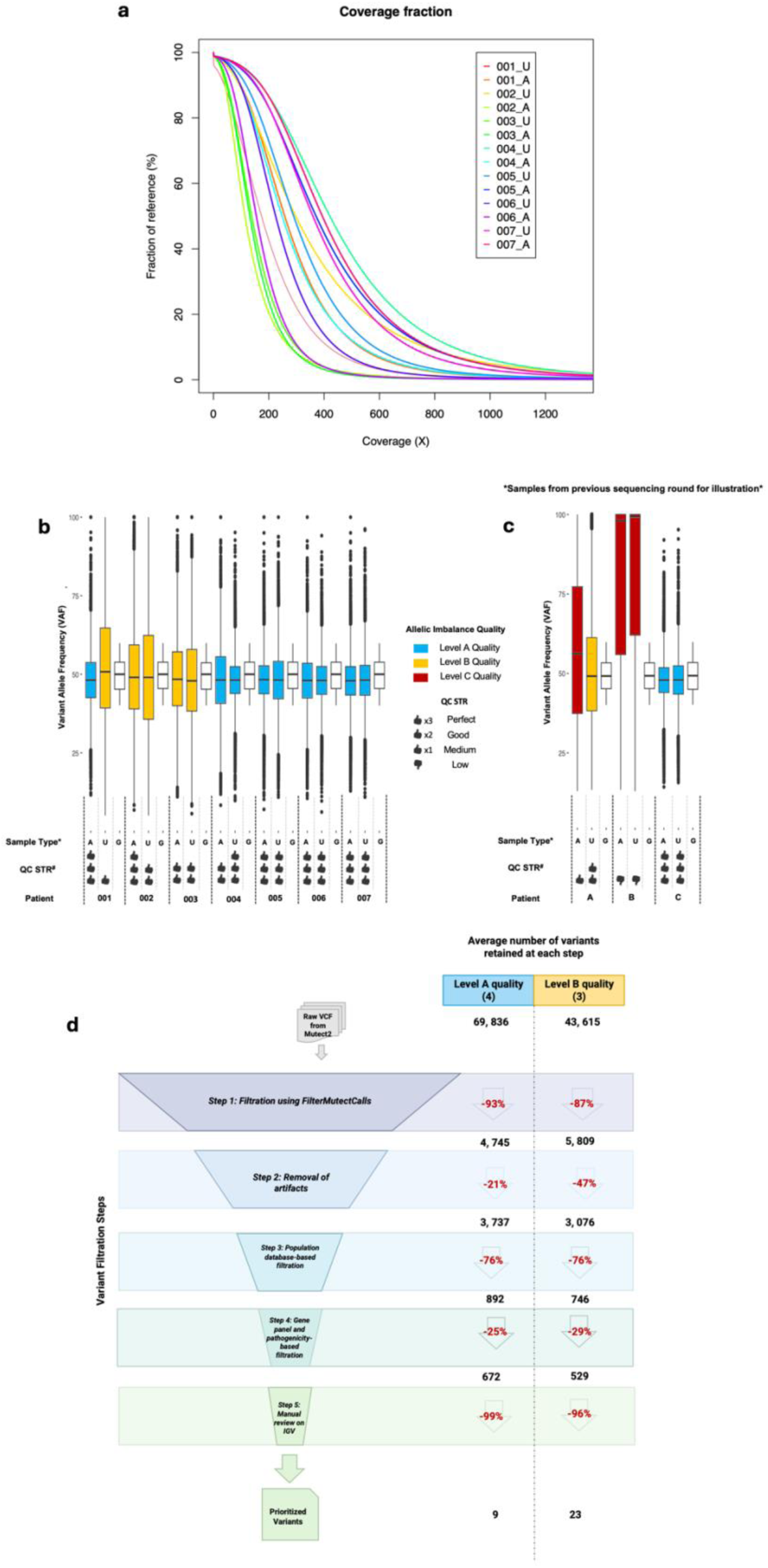
Post-sequencing quality control measures and variant filtration workflow. a, Coverage fraction: this figure illustrates which fraction of the exome (reference) was covered at which sequencing depth (coverage) for the sequenced SEEG-derived neuronal DNA samples. A = Affected brain region, U = Unaffected brain region. **b and c,** Analysis of allelic imbalance in SEEG-derived neuronal DNA and germline-derived DNA samples: This figure presents the distribution of variant allele frequency (VAF) among heterozygous variants identified in SEEG- derived neuronal DNA samples (affected and unaffected) compared with germline-derived DNA samples (blood/saliva). **b,** displays data from seven patients included in this publication. **c,** showcases data from three patients from an earlier sequencing round, before optimizing our sEEG harvesting protocol, to demonstrate the allelic imbalance quality in samples with suboptimal short tandem repeat (STR) quality. A = Affected SEEG-derived neuronal DNA samples, U = Unaffected SEEG-derived neuronal DNA samples, and G = Germline-derived DNA samples. The allelic imbalance quality is classified as follows: ’Level A’ for patient samples with affected and unaffected neuronal samples falling below the 1x SD threshold of 11.5%, ‘Level B’ for patient samples with unaffected and/or affected in between the 1x and 2x SD threshold (11.5%-23.0%), and ’Level C’ for patient samples with unaffected and/or affected exceeding this 2x the SD threshold (23.0%). The STR analysis is categorized as ’Perfect’ (no allelic dropout), ’Good’ (≤ 2 markers with dropout), ’Medium’ (≤ 4 markers with dropout), and ’Low’ (> 4 markers with dropout). These visualizations were generated using the ggplot2 package in R. **d,** Schematic illustrating the step-wise filtration process of variants from raw VCF files generated by Mutect2 and the corresponding average number of variants retained at each stage for ‘Level A’ and ‘Level B’ quality samples. Starting with raw variant calls, the process undergoes successive refinement through five major steps: 1) Filtration using FilterMutectCalls: variants with “PASS” flag were retained, 2) Artifact removal: based on in-house database, variants in <2 other neuronal samples were retained, 3) population database-based filtration: variants in <1% population frequency on GnomADv2 were retained, 4) gene panel and pathogenicity-based filtration: variants in epilepsy genes and /or variants with >12 CADD score were retained, and 5) manual review using Integrative Genome Viewer (IGV) (see methods for details), culminating in the selection of prioritized variants. The average number of variants retained after each step is displayed (in black font), alongside the average percentage reduction observed during each filtration step (in red font).

### Allelic imbalance in heterozygous variants can be used to determine sample quality prior to variant detection

During our examination of germline variants in neuronal and germline (blood/saliva) samples, we noted instances of allelic imbalance that could have affected sample quality for variant detection. Typically, heterozygous germline variants exhibit a variant allele frequency (VAF) of ∼0.5; however, in affected and unaffected SEEG-derived neuronal DNA samples with low sequencing quality we noted deviations from this frequency. Such allelic imbalances— where one allele is preferentially amplified or sequenced—pose significant challenges for somatic variant detection, introducing potential errors such as false positives or negatives, or skewed variant frequencies.

To better characterize such samples, we assessed variants from each patient’s germline sample that displayed a VAF between 40% and 60%, indicating heterozygosity. We then evaluated the VAFs of these variants in the corresponding unaffected and affected SEEG-derived neuronal DNA samples as a quality metric. The standard deviation (SD) of the VAF of the heterozygous variants was calculated for each SEEG-derived neuronal DNA sample as a reflection of the deviation from the expected 50%.

To establish a quality threshold, we analysed 202 unrelated germline exomes (see methods) and determined a threshold of 11.5%. Hence, samples with both affected and unaffected SEEG-derived neuronal DNA with SD ≤11.5% were considered ’Level A’ quality, indicating minimal allelic imbalance and reliable variant detection. Conversely, samples with affected and/or unaffected SEEG-derived neuronal DNA SD values ≥23.0% (2x threshold) were flagged as ’Level C’ quality, indicating significant allelic imbalance and potentially unreliable variant detection. Samples with affected and/or unaffected SEEG-derived neuronal DNA SD values falling between these thresholds (>11.5% to 23%) were categorized as ’Level B’ quality, indicating some degree of allelic imbalance but potentially acceptable for variant detection with additional scrutiny.

Fig. 2b shows the distribution of VAFs for heterozygous variants across the different SEEG- derived neuronal DNA samples in correlation with the STR analysis. The resulting VAF distribution predominantly fell within the expected 40%-60% range. Of these, 4/7 patient samples were determined to be of ‘Level A’ quality, and 3/7 were classified as ‘Level B’ quality based on our allelic imbalance criteria (Fig. 2b). Our analysis of a previous sequencing round (samples obtained before optimizing protocol) involving three patients indicates that samples with suboptimal STR quality typically fall into the ’Level C’ quality category (Fig. 2c).

### Prioritization of somatic variants in SEEG-derived neuronal DNA

Variant calling tools such as Mutect2 yield a high number of variants. We used a variant filtration workflow in our efforts to eliminate false positives and artifacts to select candidate variants for validation (See Methods, ‘Variant filtration for manual review’). The filtration workflow is outlined in Fig. 2d with the retention rates at each step for ‘Level A’ and ‘Level B’ quality samples. A detailed table for all seven patient samples is shown in Supplementary Tables 3 and 4.

Comparing the ’Level B’ (3/7) and ’Level A’ quality samples (4/7), we observed some differences in the number of variants retained and the percentage of reduction in variants at Steps 1 and 2 of the variant filtration workflow. At Step 1, ’Level B’ quality samples had more variants with the “PASS” flag on average (5,809) compared to ‘Level A’ (4,745), suggesting a potential overrepresentation of variants in ’Level B’ quality samples that could include false positives.

After the removal of artifacts (Step 2), ’Level A’ quality samples retained more variants (79%) than ’Level B’ quality samples (53%). This implies a better accuracy of variant calls in ’Level A’ quality samples after initial quality control, and that ‘Level B’ quality samples have a higher initial burden of artifactual variants. Further investigation showed that ‘Level B’ quality samples contained 77-416 variants called in other germline samples, while this number was lower (4-15) in ‘Level A’ quality samples.

After manual review on Integrative Genome Viewer (IGV; Step 5), ’Level B’ quality samples yielded 23 prioritized variants on average whereas ’Level A’ quality samples yielded 9 prioritized variants. From these, we selected four candidate somatic variants for digital droplet polymerase chain reaction (ddPCR) validation across two ‘Level B’ quality (#001, #002) and two ‘Level A’ quality (#005, #006) patient samples based on gene association and pathogenicity scores (Table 2).

**Table 2:** ***Candidate somatic variants for ddPCR validation.*** * This variant was previously reported^16^. Abbreviations: CADD, Combined Annotation Dependent Depletion; ddPCR, digital droplet Polymerase Chain Reaction.

| Patient ID | Number of neuronal nuclei in affected sample | Chrom | Position | Gene | Reference Nucleotide | Alternate Nucleotide | Type of variant | CADD Scores | VAF in exome (%) | VAF in ddPCR (%) | Detection Threshold of ddPCR (%) |
| --- | --- | --- | --- | --- | --- | --- | --- | --- | --- | --- | --- |
| #001 | 228 | 1 | 115,272,929 | CSDE1 | C | A | missense | 24.9 | 4% | 1.34% | 0.0147% |
| #002 | 250 | 17 | 33,460,232 | NLE1 | C | G | missense | 27.8 | 11% | 12.6% | 0.025% |
| #005 | 250 | 10 | 89,622,242 | KLLN | C | T | start loss | 17.3 | 4% | 1.73% | 0.173% |
| #006 | 140 | 1 | 11,301,686 | MTOR* | G | T | missense | 23.8 | 4% | 0.78% | 0.05% |

### Validation of candidate somatic variants using ddPCR

Using ddPCR, we successfully confirmed the presence of all four candidate somatic variants in their respective SEEG-derived neuronal DNA samples (Table 2, Fig. 3). These variants were absent in both unaffected SEEG-derived neuronal DNA and germline-derived DNA samples, indicating their somatic origin and association with the affected tissue (Extended Fig. 1).

**Fig. 3:**
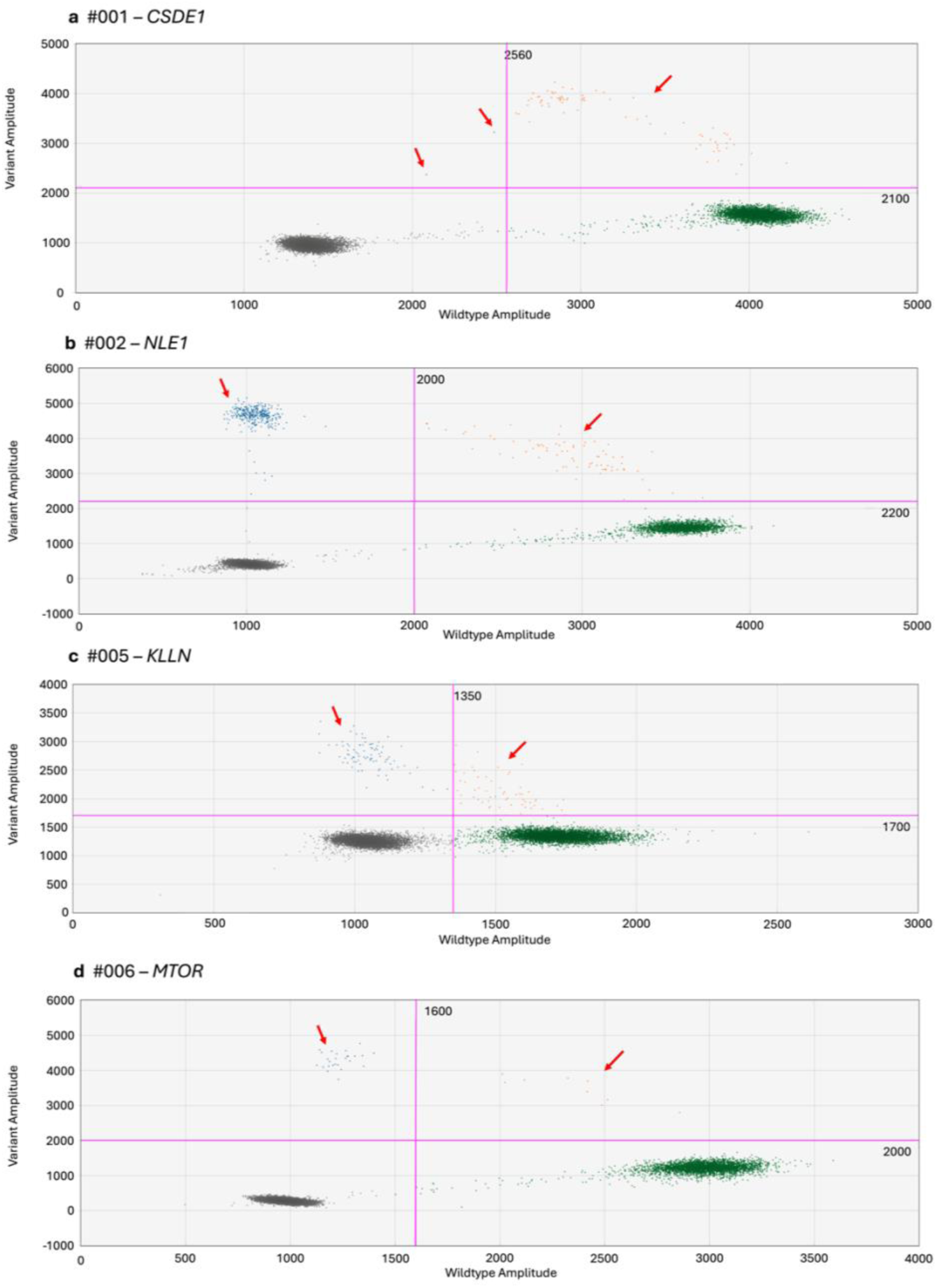
ddPCR results for candidate variants in affected brain regions showing variant confirmation in affected samples (a-d). Due to technical issues (GC-rich region), the KLLN assay did separate less well between positive and negative variant droplets. Gating was determined based on fluorescent intensities of no template control samples. Blue: variant droplets, green: wild-type droplets, orange: droplets containing multiple DNA templates, gray: empty droplets, red arrows are showing bulk of variant droplets for CSDE1, NLE1, KLLN and MTOR. ddPCR results for candidate variants in unaffected brain regions and germline samples are available as Extended Data Fig. 1.

## Discussion

Here, we describe a protocol for extracting neuronal DNA from SEEG electrodes, quality control measures pre- and post-exome sequencing, and a variant filtration workflow to detect somatic brain variants. Our protocol significantly improves upon previous methods^12–15^ by introducing nuclei isolation and sorting techniques that purify neuronal nuclei, thereby enhancing the detection of low VAF variants.

Our nuclei yield shows that only 6% of SEEG depth electrode-derived nuclei are neuronal. About 91% did not stain as neuronal, astrocyte or microglia. These nuclei could be present due to contamination and/or nuclei from degraded brain cells. Excluding these cells from downstream steps is crucial to increase the concentration of the cell type carrying the variant of interest and increase the likelihood to detect low VAF variants. Our methodology details the nuclei isolation, staining, sorting and cleanup procedures that ensure the integrity of the neuronal nuclei and their DNA. Furthermore, we used a novel DNA amplification method, PTA, which results in a more even amplification across the genome and reduces errors during the amplification process in comparison to the standard whole genome amplification process^17^.

While the collection and processing of SEEG depth electrode samples can pose challenges, we found STR analysis to be an effective means to discern and discard low-quality samples before embarking on extensive genotyping. Presence of allelic dropout, even in a single STR marker, was indicative of suboptimal exome coverage and the potential for allelic imbalance, which can present challenges in accurate variant detection. In most instances, the STR analysis provided a reliable predictor of exome quality and the likelihood of allelic imbalance, affirming its use as a quality control measure prior to sequencing. However, limitations of STR analysis were observed, such as its ineffectiveness in detecting allelic dropout in homozygous markers and its limited genomic coverage, which could miss dropout events outside the analyzed regions. For example, the affected SEEG-derived neuronal DNA sample from #002 showed no allelic dropout in STR analysis yet exhibited signs of somewhat reduced sample quality in subsequent exome coverage and allelic imbalance assessments.

The examination of allelic imbalance at heterozygous sites post-sequencing provided further insight into sample integrity to determine the reliability of variant detection for each sample. We observed that ‘Level B’ quality samples contained a higher proportion of artifactual and false positive variants in comparison to ‘Level A’ quality samples. Consequently, a greater number of variants needed to be scrutinized to identify the true positives in ‘Level B’ quality samples. Our variant filtration workflow serves as a robust framework for the reduction of artifactual variants, increasing the possibility that only relevant and reliable somatic variants called using Mutect2 are subjected to manual review.

Using ddPCR, we validated 4/4 somatic variants chosen based on gene association and pathogenicity scores in SEEG-derived neuronal DNA from the affected brain regions and confirmed their absence in the respective SEEG-derived neuronal DNA from unaffected tissue and germline sample. Among the identified variants, one was in *MTOR*^21^, which is known to be implicated in FCD^2,4,5,8,10,22,23^. We identified a variant in *CSDE1,* which is associated with epilepsy^24^. Another variant was identified in *KLLN,* which is related to Cowden syndrome, linking it to neurological manifestations^25^. We also identified a variant in *NLE1*, which while less clearly associated with epilepsy, may represent a novel candidate gene for focal epilepsy. The validated somatic variants had VAFs between 0.78-12.6%, which corresponds to 1.56-25.2% of neurons in the affected brain regions.

We tested two widely used somatic variant callers, Strelka2 and Mutect2^10,13,19,26–28^. Strelka2 yielded ∼80% more variants than Mutect2 after initial filtering when retaining only variants with “PASS” flag. However, upon examination of these variants on IGV, the majority of these calls were of low quality (lower <20 reads coverage in either of the affected, unaffected or germline samples, low quality reads, etc.) This led us to choose Mutect2 in our workflow over Strelka2.

Overall, the methodological improvements through the introduction of neuronal nuclei sorting and use of PTA for DNA amplification enhance the precision and reliability of our variant detection analysis. Our results also show the utility of using STR analysis as a quality control measure before sequencing. In addition, the allelic imbalance analysis we use informed us about the reliability of the sequenced patient samples, allowing us to be cautious with variant assessment in some samples.

### Limitations

Our approach uses strict quality control measures to select high quality samples for further analysis. Out of 17 patients from whom SEEG electrodes were collected, 9 fulfilled our quality criteria. Hence, it is expected that about 45% of the samples will not result in high quality DNA. A certain percentage of these could probably still be analyzed but exclusion of artifacts will require more effort during filtering. Future work will be required to determine the exact threshold for further analysis of SEEG-derived DNA.

A potential limitation of the SEEG harvesting method is the sensitivity to identify variants with low VAF. As our technique provides the exact number of nuclei of each cell type that are being amplified ^21^, it allows estimation of the power to identify a certain VAF. 80% of the pools included ≥112 neuronal nuclei corresponding to 80% power to identify a VAF of ≥0.7%. Hence, the majority of samples provides high sensitivity to identify low VAF.

The variants presented here were identified in neuronal nuclei. However non-neuronal cells have also been implicated in epileptogenesis^21^. With our approach, the implication of somatic variants in other cell types such as astrocytes and microglia can also be studied. Our method can be also used with whole genome sequencing, which can uncover non-coding and structural variants. Ultimately, our methodology contributes to a more accurate identification of somatic variants in SEEG-derived DNA, paving the path to understanding the molecular basis of epilepsy as well as improving diagnosis, therapeutics and precision medicine strategies. Our method is adaptable and can be applied to any context where small amounts of tissue are collected, where there is a potential for contamination, which may include biopsies in both human and animal studies.

## Detailed Protocol

### SEEG depth electrode samples

SEEG depth electrodes were obtained during explantation from adult and pediatric patients undergoing an SEEG procedure. The tips of the depth electrodes including all contacts were collected in 15 ml Falcon® tubes containing ∼7 ml precooled PBS, and they were transported on ice. Electrodes were combined by region aiming to include 2-4 electrodes per region with one region representing the seizure onset zone and one other region an unaffected cortical area. Metal bolts were used in the majority of patients. During collection, the electrodes were severed below the bolt to avoid contamination as the bolts are exposed to the outside environment and may contain skin and/hair. In some cases, the electrodes were removed before the bolts (i.e. pulled through the bolts). In these cases, the electrodes were severed right above the last metal contact. The electrodes were placed in the Falcon® tubes immediately to avoid DNA degradation due to prolonged exposure to air. The depth electrodes in the Falcon® tubes were transported on ice and stored at -80°C within 10-15 minutes after collection was completed to maintain DNA quality.

The study was approved by the Conjoint Health Research Ethics Board (CHREB) at the University of Calgary (REB18-2099). The patients or their legal guardians provided written informed consent.

### Blood and saliva-derived DNA samples

Blood or saliva samples from patients were obtained for comparison. DNA was extracted using standard methods.

### Preparation of FANS buffer

FANS buffer was used to maintain the integrity of the isolated nuclei during the nuclei isolation procedure. The FANS buffer was prepared by adding 5 ml of 10% BSA (Bovine serum albumin) and 100 *µ*l of 0.5 M EDTA to 45 ml of DPBS (Dulbecco’s phosphate-buffered saline), following the protocol outlined in *Nott et al. 2021* ^18^. The prepared FANS buffer was stored at 4°C for up to a week. For each sample, approximately 3 ml of FANS buffer was required during the nuclei isolation and staining procedure.

### Preparation of antibody stains

To purify neuronal nuclei, isolated nuclei were stained with antibodies. DAPI stain was used to select nuclei containing DNA. 0.5mg/ml DAPI working stock solution was prepared according to *Nott et al.*^18^. The working stock solutions for anti-NeuN and anti-SPI1 antibodies were prepared as follows: 5 µl of the anti-NeuN antibody (Abcam; ab190195) was added to 45 *µ*l of PBS+5%BSA. 5 µl of anti-SPI1 antibody (Biolegend; 658010) was added to 45 µl of PBS+5%BSA. All antibody stock solutions were kept protected from light, and the working stock solutions of DAPI, Anti-NeuN and Anti-SPI1 were stored at -20°C. The working stock solution for anti-LHX2 (USBiological; 129079-ML650) was prepared as needed and kept at 4°C because it cannot be stored long term. To prepare the antibody cocktail for the nuclei staining process, the required amount of each antibody was mixed according to the number of samples being processed (Table 3). Each sample required 8 µl of the antibody cocktail.

**Table 3:**
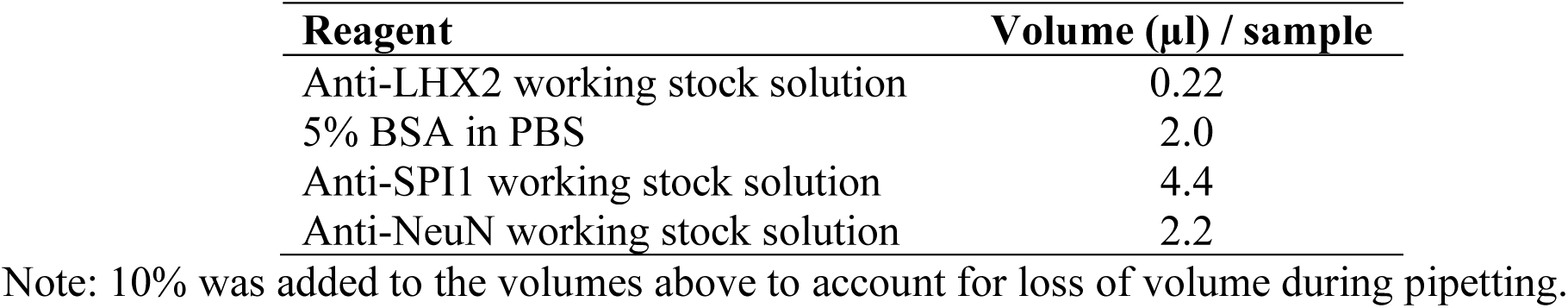
Antibody cocktail preparation.

| Reagent | Volume (μl) / sample |
| --- | --- |
| Anti-LHX2 working stock solution | 0.22 |
| 5% BSA in PBS | 2.0 |
| Anti-SPI1 working stock solution | 4.4 |
| Anti-NeuN working stock solution | 2.2 |

Note: 10% was added to the volumes above to account for loss of volume during pipetting.

## Procedure

### Cell harvesting from depth electrodes

Samples of frozen depth electrodes in PBS stored at -80°C in Falcon® tubes were thawed at room temperature until the outer parts of the sample started to melt. The tubes were then transferred to a beaker containing ice and placed on a nutating shaker for approximately one hour to maintain a uniform low temperature of the liquified portion of the sample and facilitate cell release from the depth electrodes.

After complete thawing, the tubes were vortexed at 1200 RPM several times to aid in the release of cell from the depth electrodes. Using sterile tweezers, the depth electrodes were carefully removed from the tube. The Falcon® tubes containing the released cells were centrifuged at 800g for 10 minutes in a cooled centrifuge.

Following centrifugation, the supernatant was carefully decanted, leaving behind approximately 300 µl of volume in the tube. Note: A pellet is usually not present.

### Nuclei isolation

Nuclei were isolated using the Minute^TM^ Single Nucleus Isolation Kit for Neuronal Tissues/Cells (Invent Biotechnologies BN-020). As larger debris was typically absent in our samples, the use of the supplied filter was omitted for maximum recovery of nuclei. 200 µl of cold buffer A was added to each sample and vortexed at 1200 RPM. If a visible pellet remained, the sample was pipetted up and down using a 20-100 µl pipette until this was resuspended. The walls of the tubes were washed using an additional 400 µl of cold buffer A to ensure that any cells adhering to them were properly dislodged. The samples were then vortexed again at 1200 RPM and incubated at -20°C for 15 minutes, with caps open. Following the incubation period, the samples were centrifuged at 800 g for 5 minutes. The supernatant was subsequently discarded, leaving behind an approximate volume of 300 µl containing the isolated nuclei.

The isolated nuclei were subjected to a clean-up step to eliminate any cellular debris, oil and myelin. For each sample, a new 1.5 ml Eppendorf tube, pre-labelled with sample ID and brain region and containing1 ml cold buffer B (kit BN-20) was prepared. Following this, 150 µl of cold PBS containing 5% BSA was added to resuspend the isolated nuclei sample, which was subsequently overlaid onto the buffer B by carefully dispensing the liquid against the Eppendorf tube wall. To ensure no nuclei remained in the Falcon tube, an additional 50 µl of cold PBS with 5% BSA was used to rinse the walls of the Falcon tube, and the volume was also transferred to the Eppendorf tube and overlaid onto the buffer B. The Eppendorf tubes containing the isolated nuclei samples and buffer B were centrifuged at 1500 g for 10 minutes. During the centrifugation, the isolated nuclei pass through buffer B and settle at the bottom of the Eppendorf tube whereas contaminants may form a milky layer on top of buffer B which was carefully eliminated. If no visible milky layer was observed, approximately 1.2 ml of the volume was removed from the top, leaving approximately 20-25 µl of volume containing the isolated nuclei. Subsequently, 500 µl of FANS buffer was added to resuspend the isolated nuclei, and this volume was transferred to 15 ml Falcon tubes. An additional 1 ml of FANS buffer was used to rinse the wall of each Eppendorf tube and transferred again to the respective Falcon tube.

Finally, the samples were vortexed at 1200 RPM.

Control samples

To determine the level of background fluorescence, an unstained sample was prepared using 25 µl of one sample which was transferred to a new Falcon tube with 75 µl of FANS buffer. This sample was labelled the unstained control.

Additional 25 µl of one sample were transferred to a separate Falcon tube and combined with 75 µl of FANS buffer as DAPI control sample.

Single stain controls were prepared for anti-NeuN, anti-SPI1 and anti-LHX2 in three separate tubes. 20 µl of UltraComp eBeads™ Plus Compensation Beads (Invitrogen: 01-3333- 41) were added to each tube after they were mixed by pulse vortexing. To each tube the respective antibody working stock solution was added: 2 µl of anti-NeuN working stock solution, 4 µl of anti-SPI1 working stock solution and 2 µl of anti-LHX2 working stock solution. The three tubes were then incubated at room temperature in the dark for 15 minutes. Following incubation, 3 ml of FANS buffer was added to each tube and centrifuged at 400-600 g for 3-5 minutes. The supernatant was decanted. 300 µl of FANS buffer was added to each tube and mixed using pulse vortexing.

Nuclei staining

The samples were centrifuged at 800 g for 10 minutes followed by careful removal of the supernatant leaving behind ∼100 µl in each tube. Subsequently, 8 µl of the antibody cocktail was added to each sample and vortexed. The tubes containing the samples with the antibody were covered with aluminium foil and incubated for 1 hour on ice on a nutating shaker.

Subsequently, 1.5 ml of FANS buffer was added into each Falcon® tube containing the antibody-stained samples and centrifuged at 800 g for 10minutes. The supernatant was then discarded leaving behind a volume of 200 µl. Next, 2 µl of 0.5mg/ml DAPI working stock solution was added to each antibody-stained sample, whereas 1 µl of 0.5mg/ml DAPI working stock solution was added to the DAPI control sample. The samples and DAPI control were incubated on ice, covered with aluminium foil for 10 minutes. After the incubation, the samples were subjected to FANS.

### Nuclei sorting

The gating strategy determines how nuclei populations are chosen during sorting for downstream analysis (Extended data Fig. 2). First, nuclei were selected based on granularity and size. DAPI positive nuclei were then gated based on the threshold determined by the fluorescence in the DAPI control and the autofluorescence in the unstained control. Based on the fluorescence of the positive antibody controls and autofluorescence in the unstained sample, the population of DAPI positive nuclei was then further gated into neuronal (DAPI+, NeuN+, SPI1-), astrocyte (DAPI+, LHX+, NeuN-, SPI1-), microglia (DAPI+, SPI1+, NeuN-) and other nuclei (DAPI+, NeuN-, LHX2-, SPI1-).

The antibody-stained samples were sorted with FANS and neuronal nuclei were collected in Eppendorf tubes containing 30 µl of 1.5% BSA to prevent clumping and to maintain the integrity of the nuclei. To avoid variability in the number of nuclei in the aliquoted sorted nuclei samples, the neuronal nuclei were divided into five tubes based on the count of the sorter: two tubes containing 250 nuclei, two tubes containing 500 nuclei, and one tube with the remaining neuronal nuclei. For the first tube, the yield precision mode was used to maximize the collection of neuronal nuclei, particularly in the instances when there was a low number of neuronal nuclei present in the sample. For the remaining tubes, the purity precision mode was utilized to ensure the reduction of non-target particles in the sorted nuclei samples.

### Nuclei cleanup

After nuclei sorting, the neuronal nuclei underwent a series of washing and reduction steps to minimize the BSA concentration prior to whole genome amplification. The Eppendorf tubes harboring the sorted nuclei were washed with 60 µl ResolveDNA PTA-Grade cell buffer (BioSkryb Genomics 100002) to dislodge any nuclei that may adhere to the walls of the tube. Subsequently, the tubes were centrifuged at 100 g for 30 seconds. The resulting volume was transferred to a PCR tube and the walls of the Eppendorf tube were then washed with an additional 60 µl ResolveDNA PTA-Grade cell buffer which was also transferred. The PCR tubes were then subjected to a 10-minute centrifugation at 800 g and, thereafter, the volume in the PCR tube was reduced to 6 µl via pipetting. An additional 60 µl ResolveDNA PTA-Grade cell buffer was added to the reduced nuclei samples, which were then centrifuged again at 800 g for 10 minutes. The final volume was once again reduced to 6 µl using a pipette. Finally, the nuclei samples were appropriately labelled and stored for short-term at -20°C or for long-term storage at -80°C.

### Whole genome amplification

Whole genome amplification was performed from purified neuronal nuclei of the affected region as well as from purified neuronal nuclei from a clearly unaffected region. Depending on the nuclei yield, multiple neuronal nuclei aliquots from the same region were available.

Amplification was performed on nuclei samples sorted using purity precision mode whenever possible; otherwise, nuclei samples sorted using yield precision mode were used (see “Nuclei Sorting” above).

Neuronal nuclei samples (6 µl) were thawed and subjected to DNA amplification using PTA (BioSkryb ResolveDNA Whole Genome Amplification Kit, 100372), with two modifications to the protocol provided. Specifically, all reagents were doubled in volume to maximize DNA yield. This ensured that sufficient reagents were available for proper amplification to occur and reduced the concentration of possible inhibiting contaminants (such as low molecular DNA) in the sample. We also increased the incubation time during the lysis step from 1 to 20 minutes to improve nuclei lysis as recommended by BioSkryb.

After DNA amplification, we purified the products using the ResolveDNA Bead Purification Kit (100182). Although the amplification involved a double reaction, which typically necessitates doubling the purification reagents, we observed higher DNA yields when processing half of the volume of the amplified DNA for a single purification reaction.

Consequently, we opted to purify half of the amplified neuronal samples with a single reaction to enhance DNA recovery. The remaining sample was stored at -80°C for future cleanup. DNA yield of amplified samples, positive and negative control were quantified using Qubit^TM^ fluorometer using the Qubit^TM^ dsDNA Quantification HS Assay kit (ThermoFisher Scientific, Q32854).

### Pre-sequencing quality control

Purified neuronal DNA samples and the corresponding blood- or saliva-derived DNA were subjected to analysis of multiple STR markers (D21S11, CSF1PO, vWA, D8S1179, TH01, D18S51, D5S818, D16S539, D3S1358, D2S1338, TPOX, FGA, D7S820, D13S317, D19S433) to confirm genome-wide amplification and exclude DNA contamination during the amplification process. The AmpFLSTR^TM^ Identiflier^TM^ Plus (Applied Biosystems^TM^, 4427368) PCR amplification kit was used for this purpose according to manufacturer’s protocol. The purified neuronal DNA samples were also subjected to automated electrophoresis using the TapeStation D5000 assay to ensure the presence of DNA between 200-4000 bp as expected for amplification with the PTA method. High quality samples were then selected for whole exome sequencing.

### Exome sequencing

For each patient, one DNA sample derived from neuronal nuclei from one affected region, one unaffected region and germline (blood/saliva) was subjected to sequencing. A total of 14 SEEG-derived neuronal DNA samples (7 affected regions and 7 unaffected regions) underwent sequencing, accounting for 90% of the pooled material. Additionally, the corresponding germline (blood/saliva derived DNA) samples underwent a single capture, constituting the remaining 10% of the pooled material. The exome capture was performed using the IDT xGen Human Exome V2 panel. The sequencing process was executed on the NovaSeq 6000 S1 platform, employing a 2x100 bp flow cell type (Centre for Health Genomics and Informatics at the University of Calgary). The pooling strategy was designed to achieve different coverage depths for the two capture groups, aiming for approximately 400x coverage per sample in the 90% pool (SEEG-derived neuronal DNA samples), and around 90x coverage per sample in the 10% pool (blood/saliva-derived DNA samples).

### Alignment

The generated FASTQ files (sequence files) underwent pre-alignment quality control using FastQC and trimming of low-quality bases using Trimmomatic^26^. BWA-MEM^27^ was used to perform sequence alignment against the human reference genome, resulting in bam files (alignment files) ^28,29^.

To ensure robustness of the post-alignment quality assessment, Qualimap was used to obtain an overview on mean exome coverage and fraction of the exome covered by sequencing^20^. Qualimap analysis was performed on bam files corresponding to samples from each patient.

Moreover, MultiQC was implemented to comprehensively aggregate post-alignment quality metrics across multiple samples^30^.

### Allelic imbalance analysis

To determine the quality threshold, germline variants were called (as in ^28,29^). on 202 unrelated germline (blood/saliva) samples obtained from patients with epilepsy for another project Variants called in blood or saliva samples were filtered to only include variants with VAF > 20% and <80%. This range was selected to include as much of the distribution around 0.5 (heterozygous variants) but exclude the peak at 1.0 (homozygous variants). For these variants, SD of the VAFs was calculated (11.5%) and set as quality threshold.

To evaluate allelic imbalance, germline variant calling was conducted on blood/saliva, affected neuronal and unaffected neuronal samples (as in ^28,29^). Variants called in blood or saliva samples were filtered to only include definite heterozygous variants (VAF > 0.4 and <0.6). The VAF of the heterozygous variants were then assessed in the affected and unaffected SEEG-derived neuronal DNA samples using an in-house python script (available on GitHub) and SD was calculated for each sample. This SD was then compared with the established threshold from blood/saliva samples (11.5%).

Samples with both affected and unaffected SEEG-derived neuronal DNA falling below 1x SD (11.5%) were considered ’Level A’ quality. Conversely, samples with affected and/or unaffected SEEG-derived neuronal DNA falling beyond 2x SD (23.0%) above the threshold were flagged as ’Level C’ quality. Samples with affected and/or unaffected SEEG-derived neuronal DNA falling between these thresholds were categorized as ’Level B’ quality.

### Somatic variant calling

Somatic variants were called using Mutect2^31^ with default parameters, a variant calling tool specific to somatic variants, in the affected neuronal sample, with the unaffected neuronal sample specified as the matched control.

### Variant filtration for manual review

The variant filtration strategy is outlined in Fig. 2d. For variants called using Mutect2, the FilterMutectCalls tool was used to filter out sequence context-based artifacts. The tool adds a variant quality flag to the variants called, and only variants with the “PASS” flag were chosen (Fig. 2d, Step 1).

In the second filtration step, variants that were present in more than two other unaffected or affected SEEG-derived neuronal DNA samples or at least one other blood/saliva-derived DNA sample were excluded, as these likely represented amplification or sequencing artifacts (Fig. 2d, Step 2). To achieve this, an in-house database with variants called across all sequenced SEEG-derived neuronal DNA samples and blood/saliva-derived DNA samples was created. A custom python script (available on Github) was utilized to read the VCF file generated from the previous step corresponding to the affected SEEG-derived neuronal DNA sample, and each variant was cross-referenced against the in-house variant database. The resulting output VCF file included variants found in two or fewer unaffected SEEG-derived neuronal DNA samples and not found in any other affected SEEG-derived neuronal DNA sample or blood/saliva-derived DNA sample.

The resulting filtered list of variants were annotated using SnpEff ^32^ and filtered using population databases (gnomADv2^33^, TOPmedv3^34^) to exclude variants with population frequency of >=0.01 (as in ^28,29^; Fig. 2d, Step 3).

Two virtual gene panels were used for initial analysis of variants. The first gene panel consisted of 165 genes associated with the PI3K/AKT/*mTOR* pathway^35^. The second gene panel consisted of 954 genes associated with epilepsy^36^. The resulting filtered list of variants from Step 3 were flagged if present in the gene panel genes and subjected to manual review on IGV^37^ (see below). Variants in other genes (not included in the gene panels) were filtered based on pathogenicity scores (CADD 1.4 score >= 12). These variants were also manually reviewed on IGV for prioritization (Fig. 2d, Step 4).

### Manual review of somatic variants

Manual review of the variants was done on IGV^37^ to exclude artifacts and involved comparing the presence of the variant in the sequenced samples of affected SEEG-derived neuronal DNA, unaffected SEEG-derived neuronal DNA, and germline-derived DNA (Fig. 2d, Step 5). Comparison with germline (blood/saliva) was used to eliminate any germline variants erroneously identified as somatic. The unaffected SEEG-derived neuronal DNA sample was used to determine if the variant was only present in the affected SEEG-derived neuronal DNA sample, or if it was present in the unaffected SEEG-derived neuronal DNA sample at a lower frequency in comparison to the affected SEEG-derived neuronal DNA sample.

During manual review, variants were examined based on several criteria^38^. Variants were excluded if a) they were present in affected SEEG-derived neuronal DNA but also present in both unaffected and blood, b) they were present at similar frequencies in both affected and unaffected SEEG-derived neuronal DNA, c) they had less than two supporting reads (i.e. reads with the variant present) or two supporting reads that were in the same directions d) they were present in only reads in the same direction and in reads of the same size, e) they had low mapping or base quality (below the default threshold 20 for both), f) they were present at a site that was covered by less than 20 reads in the affected, unaffected and blood samples.

Variants that passed manual review in each patient sample were classified as “initial prioritized variants”. These initial prioritized variants were further manually reviewed on IGV in other germline and neuronal tissue samples that were sequenced using the same approach, to exclude any amplification or sequencing artifacts. Variants were excluded if present in more than two other unaffected or affected SEEG-derived neuronal DNA samples (19 unaffected samples), or at least one blood/saliva-derived DNA sample (10 blood/saliva samples). Doing so ensured that variants present in only a fraction of the reads that were not called by Mutect2 were also interrogated. The resulting variants were termed “prioritized variants”. Prioritized variants that passed this manual review were then queried for gene function and association with epilepsy or seizures to obtain “candidate variants” for validation.

### ddPCR validation

Droplet digital PCR (ddPCR) was performed for variant validation using a QX200 Droplet Generator (Bio-Rad), QX200 Droplet Reader (Bio-Rad) and custom TaqMan® SNP genotyping Assays (Thermo Fisher Scientific) using 60 ng of SEEG-derived affected and unaffected neuronal DNA and in blood- or saliva-derived DNA. The data was analyzed with the QX Manager Software 2.1 Standard Edition (Bio-Rad). The detection limit of the assay was established using wildtype DNA spiked with decreasing concentrations of mutant gBlock (Integrated DNA Technologies, Inc.). The primer and probe sequences, and annealing temperatures for the four variants are available in the supplementary information. As the *KLLN* variant was located in a GC-rich region, the gBlock was modified outside of the assay binding region to allow synthesis.

## Data availability

The deidentified phenotypic and exome sequencing data will be made available upon reasonable request from any qualified investigator after institution of a material transfer agreement.

## Code availability

The in-house python scripts used for the removal of artifacts and allelic imbalance analysis is made available on GitHub: https://github.com/MTG-Lab/SomaticVariantAnalysis

## Acknowledgements

We would like to thank the patients and their families for participation in the study. We acknowledge the Flow Cytometry Core Facility and the Centre for Health Genomics and Informatics at the University of Calgary for their support. This project has been made possible by the Canada Brain Research Fund (CBRF), an innovative arrangement between the Government of Canada (through Health Canada) and Brain Canada, and by the Azrieli Foundation. It was also supported by Epilepsy Canada, doing business as Epilepsy Canada. The 202 unrelated germline samples were sequenced within the RAISE-GENIC project (Rational antiepileptic drug selection by combining gene network and ICT analysis), funded by the Canadian Institutes of Health Research (CIHR), under the frame of ERA PerMed. KMK also received support from the Department of Clinical Neurosciences, Hotchkiss Brain Institute and Alberta Children’s Hospital Research Institute, University of Calgary. M.T-G was supported by the Alberta Children’s Hospital Foundation and the CIHR-Bridge grant number OGB-185746. Infrastructure in the laboratory of GP was supported by a Canada Foundation for Innovation Grant (John R. Evans Leaders Fund (CFI-JELF) 36624). The views expressed herein do not necessarily represent the views of the Minister of Health or the Government of Canada. Fig. 1b was created with Biorender.com.

## Author information

Authors and Affiliation

**Department of Clinical Neurosciences, University of Calgary, Calgary, AB, Canada**

Rumika Mascarenhas, Daria Merrikh, Maryam Khanbabaei, Navprabjot Kaur, Navid Ghaderi, Tyler Soule, J.P. Appendino, Julia Jacobs, Samuel Wiebe, Walter Hader, Gerald Pfeffer & Karl Martin Klein

Alberta Children’s Hospital Research Institute, University of Calgary, Calgary, AB, Canada

Rumika Mascarenhas, Daria Merrikh, Tatiana Maroilley, Julia Jacobs, Gerald Pfeffer, Maja Tarailo-Graovac & Karl Martin Klein

Hotchkiss Brain Institute, University of Calgary, Calgary, AB, Canada

Rumika Mascarenhas, Daria Merrikh, Tyler Soule, Julia Jacobs, Samuel Wiebe, Gerald Pfeffer & Karl Martin Klein

Department of Medical Genetics, Cumming School of Medicine, University of Calgary, Calgary, AB, Canada

Tatiana Maroilley, Gerald Pfeffer, Maja Tarailo-Graovac & Karl Martin Klein

Department of Biochemistry and Molecular Biology, Cumming School of Medicine, University of Calgary, Calgary, AB, Canada

Tatiana Maroilley & Maja Tarailo-Graovac

Flow Cytometry Core Facility, University of Calgary, Calgary, AB, Canada

Yiping Liu

Department of Pediatrics, University of Calgary, Alberta Children’s Hospital, Calgary, AB, Canada.

J.P. Appendino & Julia Jacobs

Department of Community Health Sciences, University of Calgary, Calgary, AB, Canada

Samuel Wiebe & Karl Martin Klein

O’Brien Institute for Public Health, University of Calgary, Calgary, AB, Canada.

Samuel Wiebe

Clinical Research Unit, Cumming School of Medicine, University of Calgary, Calgary, AB, Canada.

Samuel Wiebe

The Calgary Comprehensive Epilepsy Program Collaborators

Paolo Federico Colin Josephson William Murphy Andrea Salmon Shaily Singh

Guillermo Delgado Garcia Nathalie Jette

Morris Scantlebury Alice Ho

Yves Starreveld Goichiro Tamura Cesar Serrano-Almeida

Mostafa Fatehi Hassanabad

## Contribution

R.M. acquired data; analyzed the data; drafted the manuscript for intellectual content. D.M. acquired data; reviewed the manuscript for intellectual content. M.K. acquired data; reviewed the manuscript for intellectual content. N.K. acquired data; reviewed the manuscript for intellectual content. N.G. acquired data; reviewed the manuscript for intellectual content. T.M. analyzed the data; reviewed the manuscript for intellectual content. Y.L. acquired data; analyzed the data; reviewed the manuscript for intellectual content. T.S. acquired data; reviewed the manuscript for intellectual content. J.P.A. acquired data; reviewed the manuscript for intellectual content. J.J. acquired data; reviewed the manuscript for intellectual content. S.W. acquired data; reviewed the manuscript for intellectual content. W.H. acquired data; reviewed the manuscript for intellectual content. G.P. analyzed the data; reviewed the manuscript for intellectual content. M.T-G. designed and conceptualized the study; analyzed the data; drafted the manuscript for intellectual content; supervised; acquired funding. K.M.K. designed and conceptualized the study; major role in the acquisition of data; analyzed the data; drafted the manuscript for intellectual content; supervised; acquired funding.

## Corresponding Authors

Correspondence to Maja Tarailo-Graovac or Karl Martin Klein.

## Ethics declaration

The study was approved by the Conjoint Health Research Ethics Board at the University of Calgary (REB18-2099).

## Extended data

Extended Data Fig. 1: Droplet digital polymerase chain reaction results for candidate variants showing absence of variant in unaffected and blood/saliva-derived DNA samples.

*Extended Data Fig. 2: Nuclei gating strategy*.

## Supplementary information

### Supplementary Tables

Table 1: Detailed phenotypic information for patient samples submitted for WES.

Table 2: SEEG derived DNA sample information, STR quality and Exome coverage for samples subjected to WES. The coverage reported in this table is based on post-alignment quality of the sequences (Qualimap)

Table 3: Number of variants retained at each step across 7 patient samples.

Table 4: Average number of variants retained at each step across ’Level A’, and ’Level B’ quality patient samples.

Table 5: Primer and probe sequences, and annealing temperatures for validated variants

