## Supplemental figures for "Detecting somatic variants in purified brain DNA obtained from surgically implanted depth electrodes in epilepsy"

### Extended Data Figures

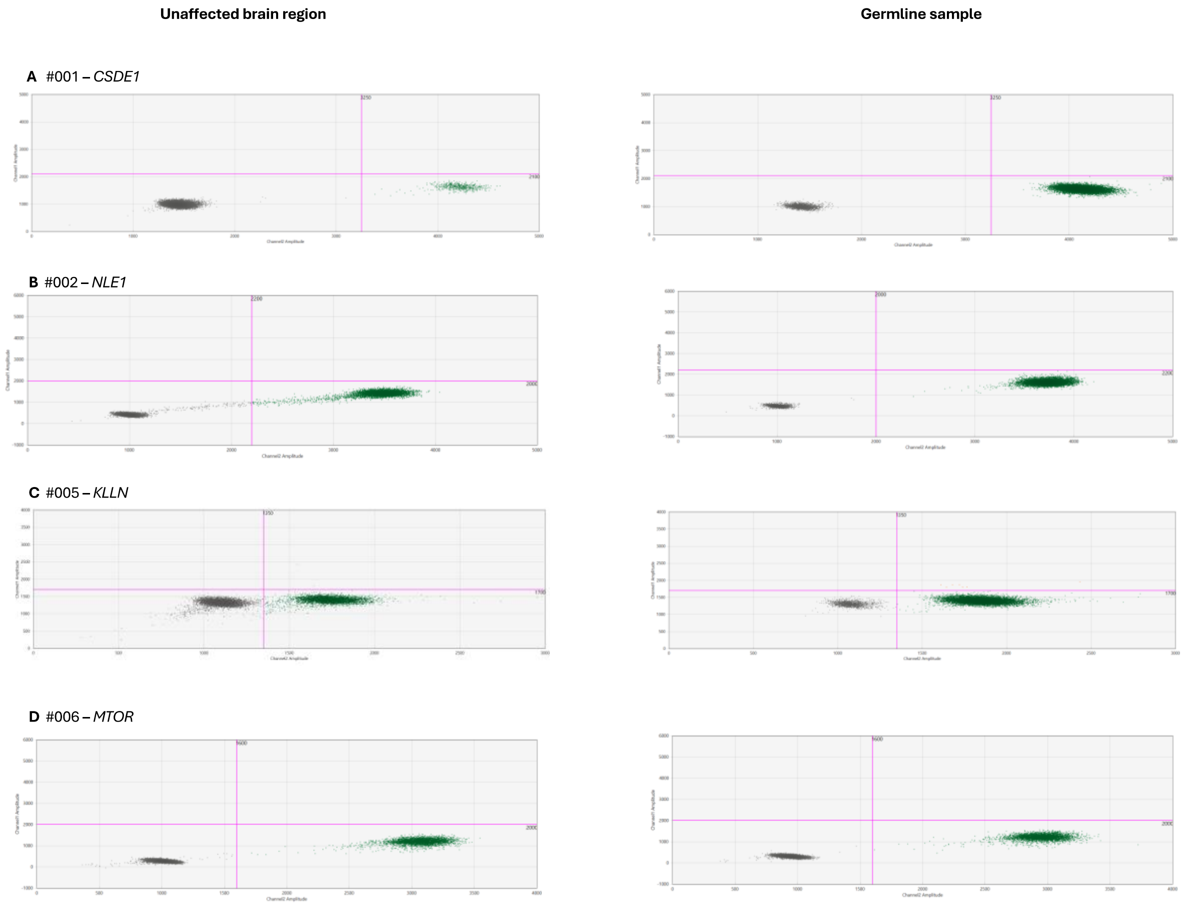


***Extended Data Fig. 1: Droplet digital polymerase chain reaction results for candidate variants showing absence of variant in unaffected and blood/saliva-derived DNA sample (A-D).*** *Due to technical issues (GC-rich region) the KLLN assay did separate less well between positive and negative variant droplets. In the unaffected neuronal sample, a small number of false positive droplets is seen which was similar to control wildtype samples (data not shown). As the number of these droplets did not exceed the determined detection threshold for this assay, these were considered false positives. Gating was determined based on fluorescent intensities of no template control samples. Blue: variant droplets, green: wild-type droplets, orange: droplets containing multiple DNA templates, gray: empty droplets.*

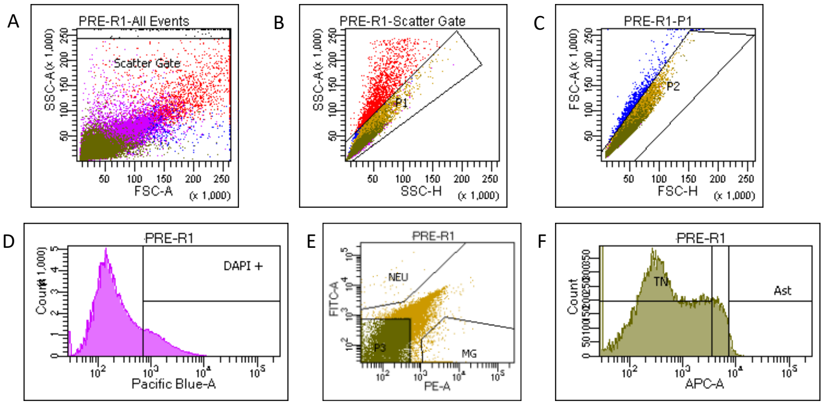


G

Tube: PRE-R1

| Population | #Events | %Parent | %Total |
|---|---|---|---|
| All Events | 244,934 | #### | 100.0 |
| Scatter Gate | 244,261 | 99.7 | 99.7 |
| P1 | 242,322 | 99.2 | 98.9 |
| P2 | 241,389 | 99.6 | 98.6 |
| DAPI + | 44,645 | 18.5 | 18.2 |
| P3 | 32,845 | 73.6 | 13.4 |
| Ast | 440 | 1.3 | 0.2 |
| TN | 26,556 | 80.9 | 10.8 |
| NEU | 45 | 0.1 | 0.0 |
| MG | 359 | 0.8 | 0.1 |

***Extended Data Fig. 2: Nuclei gating strategy. A)*** *The first gating step based on SSC-A/FSC-A aimed to identify nuclei based on granularity and size (forward vs side scatter).* ***B and C)*** *singlets were gated based on* ***B)*** *nuclei complexity or granularity P1 (SSC-A/SSC-H) and* ***C)*** *nuclei size P2 (FSC-A/FSC-H), these two gates help to exclude the doublets.* ***D)*** *The threshold for DAPI-positive nuclei was chosen based on the fluorescence in the DAPI control and autofluorescence in the unstained control.* ***E)*** *NeuN Alexa Fluor® 488 conjugated antibody and SPI1 PU.1 PE antibody were used to identify neuronal and microglial nuclei respectively. Nuclei are gated to only include those that are exclusively NeuN-positive or SPI1 positive.* ***F)*** *LHX2 APC-A conjugated antibody was used to identify astrocyte nuclei. NeuN-negative nuclei that are LHX2-positive are sorted as astrocyte nuclei. Nuclei that were negative for all three antibodies were labelled triple negative nuclei.*
